## Supplementary materials for "Multi-level analysis of adipose tissue reveals the relevance of perivascular subpopulations and an increased endothelial permeability in early-stage lipedema"

#### Supplementary information to Convolutional Neural Network (CNN)

To investigate potential differences in junction morphology between healthy/control (ctrl) and lipedema (lip) subjects, fluorescence microscopy images of endothelial cell junctions were analyzed using artificial neural networks. In particular, a multi-layer perceptron network (MLP) with an input layer, one hidden layer and an output layer was used. For our RGB images with a resolution of 250 x 250 pixels, the MLP flat input vector was 187 500 elements long. Using 10 neurons in the hidden layer and two output classes, resulted in 1 875 032 weights in the MLP network for learning. We used a small number of acceptable validation failures for evaluation patience to avoid overfitting in the learning process, which affects the classification quality. Therefore, we applied the deep learning method Convolutional Neural Network (CNN) which consists of an input layer, grouped hidden layers and an output layer. Each grouped CNN hidden layer includes a convolution layer, a rectified linear unit (ReLU) layer, a pooling layer, and optionally a normalization layer. The output layers form a fully connected layer and a classification layer (Figure S1).

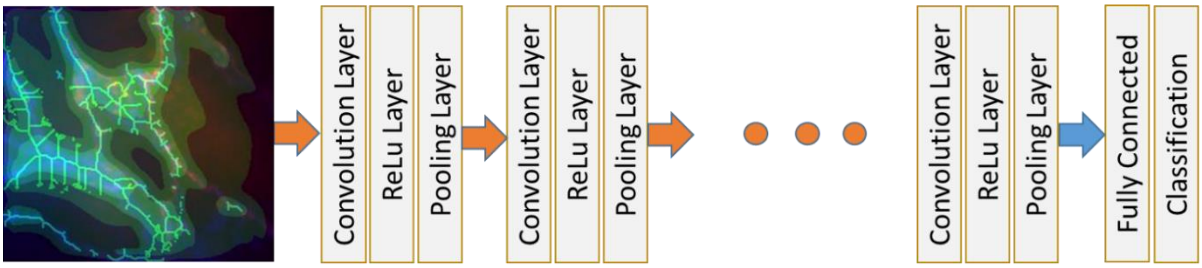


**Figure S1.** Architecture of CNN network. The CNN consists of an input layer, grouped hidden layers and an output layer. Each grouped CNN hidden layer includes a convolution layer, a rectified linear unit (ReLU)-layer, a pooling layer, and optionally a normalization layer. The output layers form a fully connected layer and a classification layer.

The CNN input data is an image matrix with the size 250 x 250 pixels for each individual red, green, and blue channel. The Convolution Layer consists of many learnable convolution filters (convolution kernels). The convolution filter, a square matrix with a small size, (e.g. 3 x 3 or 5 x 5, or n x n) is centered on an image coordinate (x, y) and then shifted from pixel to pixel over the entire image. Each corresponding output image value is calculated from the source image and the convolution filter (initialized with random numbers and modified after each iteration step of the network training) via multiplication and summation. Typically, several differently initialized filters are used to generate the outputs. These convolution output matrices are also referred to as feature maps, extracted by each filter. The result of convolutional filtering is the ReLU-Layer input, which sets the negative values of the feature maps to zero. Next, the Pooling Layer performs a sub-sampling operation, which reduces the resolution of each output features map. Usually, the mean or maximum value of the neighborhood is calculated. For example, a 2 x 2-pixel mask can be replaced by a single pixel with the maximum value in the mask. Optionally, the previously introduced layer group can be supplemented by adding a normalization layer. This complete layer group (Convolution, ReLU, Pooling and optionally

Normalization Layer) can be repeated multiple times (see Figure 1). These successive hidden layers generate generalizations of features from the preceding layer, organized in form of images. The output feature maps of the last pooling layer are converted to a single vector (flattening process) that serves as input for the Fully Connected Layer which includes neurons with softmax output functions and uses cross-entropy loss function for training. The number of neurons in the last output layer (classification layer) corresponds to the number of classes that the network should differentiate. Thus, the successive convolution layers encode the images using the learned filters and reduces the information from the vectors of all pixels to a vector of the extracted features.

The architecture of the proposed CNN network consists of 3 convolution 2D layers (Figure S2). The first convolution layer has 12 convolution filters of 3 x 3 pixels size. The first pooling layer reduces, after ReLU operation, the resolution by replacing the 2 x 2-pixel mask by one pixel with the maximum value of the mask (MaxPooling Layer). The second and third convolution layers each have 6 convolution filters with size of 3 x 3 pixels. The second and third pooling layers are identical to the first pooling layers. Before the final Fully Connected (FC) layer, a Dropout Layer was added which sets randomly a predetermined number (20%) of the input units to 0 for each step during the training time, helping to prevent overfitting.

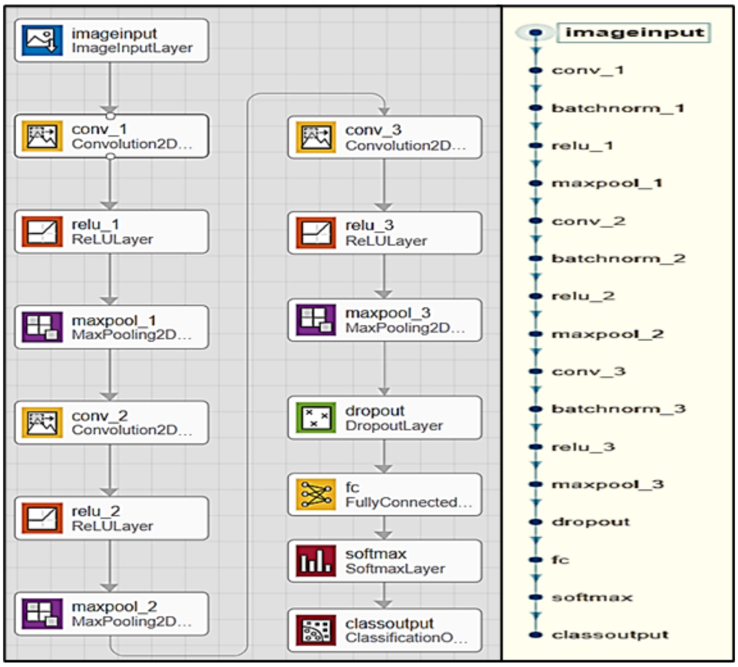


**Figure S2.** The architecture of the applied CNN network consisting of 3 convolution layers.

The trained network finally encodes input RGB-images 250 x 250 x 3 pixels (187 500 long one-dimensional vector) into 6 images with a size of 29 x 29 pixels. These final images (features maps) are transformed into a one-dimensional vector with a length of 5 048 being the fully connected layer entry.

### 1. Image-data storage and augmentation

In our study we had two sets of experiment data: 1) Stromal vascular fraction (SVF) cells were seeded to compare intrinsic junctional organization of endothelial cells (EC) from healthy (ctrl-SVF) and lipedema (lip-SVF) subjects. 2) human primary endothelial cells (hEC) were treated with SVF conditioned media (CM) from a healthy control group (ctrl-CM) and from lipedema patients (lip-CM). Each image was recorded in two color channels, the tight junction protein ZO-1 (anti-ZO-1-1A12 AlexaFluor647) in the red channel (excitation at 647 nm) and the endothelial cell marker CD31 (anti-CD31 AlexaFluor488) in the blue channel (excitation at 492 nm). For comparison, red and blue images were resized to 250 x 250 pixels. Supplmentary figure 3a depicts an exemplary, merged two-color image of a control sample of hEC. Due to the relatively small number of samples, which is typical for experiments of human tissue, we augmented the image data store by transformations of original images and combined images (saved in the green channel). In our case, additional generation of images from the originals did not distort the original image. To obtain augmented images, the original RGB image is flipped horizontally and vertically (Figure S3b and 3c, respectively).

(a) (b) (c)

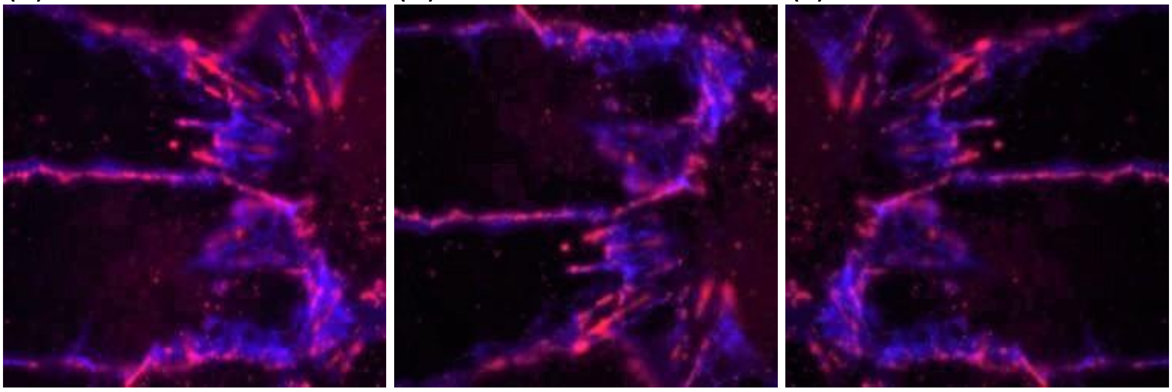

**Figure S3.** Augmentation of image data store with combined images and transformations. Cellular junctions of human primary endothelial cells were labelled with anti-ZO-1 AlexaFluor647 (red) and anti-CD31 AlexaFluor488 (blue) antibodies. (**a**) Combined original RGB image. (**b**) Augment Data: Horizontal flip. (**c**) Augment Data: Vertical flip.

Moreover, we merged the processed images of red and blue channels into the free green channel. Here, the combined image was built with filters averaging the brightness of individual pixels in relation to their immediate surroundings. This averaging corresponds to the sample density map. Both channels were filtered with a circular averaging filter (pillbox) within the square matrix of 7 pixels. Both filtered channels were then added and normalized. The resulting image was used as a green RGB-image channel (Figure S4a). This merged image was added to the augmented image data store together with its horizontal and vertical flip (Figure S4b and 4c, respectively).

(a) (b) (c)

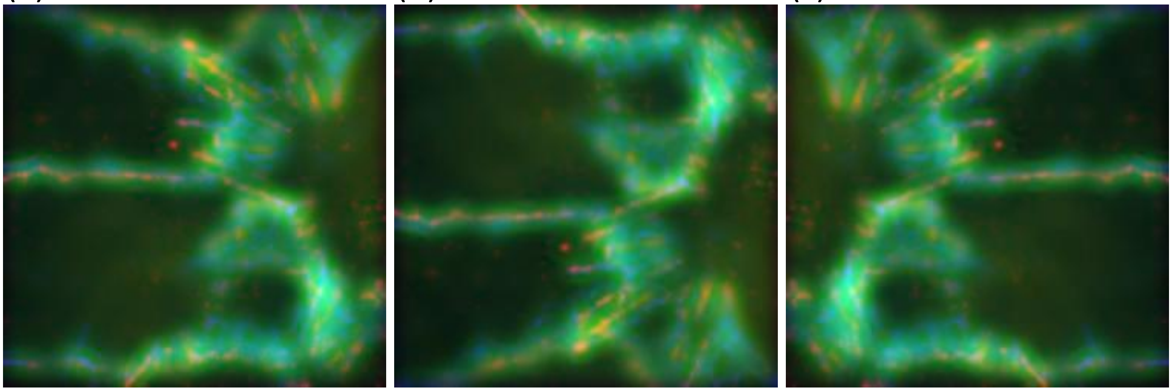

**Figure S4**. Sample density map of combined images. The combined image was built with filters averaging the brightness of individual pixels in relation to their immediate surroundings. (**a**) Density map of the combined original image. (**b**) Density map: Horizontal flip. (**c**) Density map: Vertical flip.

Additional transformations of the original image were performed using image segmentation and a skeleton operation. The segmentation of the image provides an approximate intensity map as well as the skeleton, which shows the spatial branches with high intensity. The red channel was segmented by determining 10 brightness-clusters in the image. Image brightness clustering was performed using the 2D-K-Means method. The blue channel has been binarized and skeletonized, added to the segmented red-image and normalized. This new image (Figure S5a) was used as the green channel in the new combined RGB image. This new image was also flipped horizontally (Figure S5b) as well as vertically (Figure S5c) and added to the augmented image data store. Thus, each sample/original image can be represented by a group of 9 RGB images, with each RGB image containing the original images in the red and blue channels.

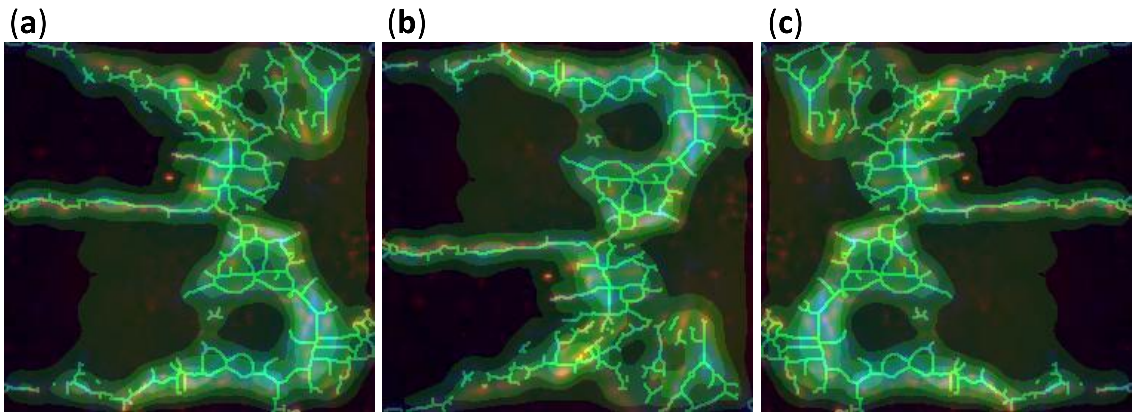


**Figure S5.** Image segmentation and skeleton operation. The image provides an approximate intensity map (mask) and the skeleton. (**a**) Mask+Skeleton RGB image. (**b**) Mask+Skeleton: Horizontal flip horizontally. (**c**) Mask+Skeleton: Vertical flip.

### 2. Experiment 1: SVF-derived EC

For the first set of data, we investigated SVF-derived EC from healthy donors (ctrl-SVF, 33 images) and lipedema patients (lip-SVF, 25 images) that were combined and augmented as described above. To prepare the test set, five samples of both groups, lip-SVF and ctrl-SVF, were selected randomly. For each of these samples, nine RGB-images with a size of 250 x 250 x 3 pixels were generated as described above. Thus, the test set contains 90 RGB images (5 x 9 images per group). The remaining 20 lip-SVF images, and 28 ctrl-SVF images formed the training set. The classes/groups were not represented equally. Such unbalanced data may make the classifier more sensitive for the class with a larger amount of data. Therefore, we generated new samples for the underrepresented class by randomly choosing samples from the currently available ones until the datasets are equalized in both training sets. Using this method, the lip-SVF group was enlarged with 8 randomly selected duplicates of the original samples, which gave 504 (252 per group) RGB-images with the size of 250 x 250 x 3 in the training set. Due to the relatively small RGB image sets, it was necessary to use a small number of acceptable validation failures (<10) for evaluation patience. Then, 10 training sessions (separately for each test and training set) of the CNN network with the previously described structure were carried out. The accuracy of the classification ranged from 76% to 96%. The average accuracy of the classification for 10 training sessions is 83.5% (± 6%). Classification accuracy of the test data (90 images) is 82.2 %. For ctrl-SVF classification accuracy is 86.7% and for lip-SVF classification accuracy is 77.8% (Figure S6).

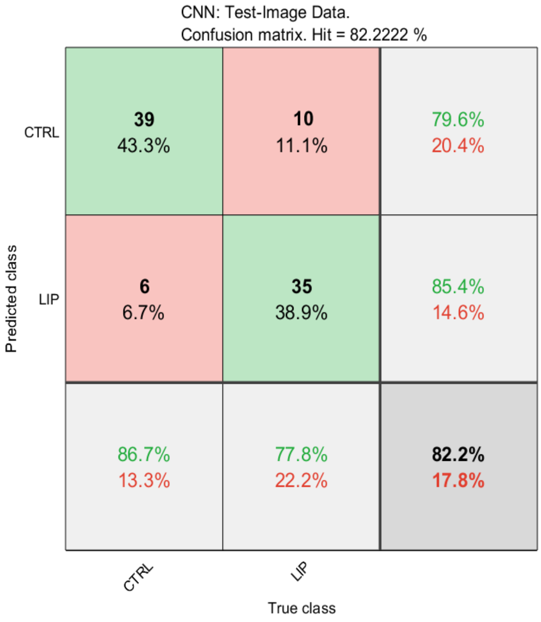

**Figure S6.** Classification accuracy shown as a confusion matrix for a test data set of experiment 1 (SVF-derived EC). The columns represent the true classes and the rows the predicted classes. The matrix diagonal green boxes give the absolute and percentage numbers of correctly classified images, and the red boxes give the amounts of false classification. The last row of the matrix (grey boxes) gives the cumulative percentage of correct (black letters) and false classifications (red letters) for both classes.

The trained CNN can be used as a classifier to predict the belonging of individual samples to a distinct class. If 9 augmented images are generated for an image and fed to the network input, the network will calculate the posteriori probability of the belonging to either the lip- or the ctrl-group. Two classification examples are shown in Figure S7. The true ctrl-SVF sample was classified as the control group with a probability of 66.7% (Figure S7a) and the true lip-SVF image was classified as the lipedema group with a probability of 74.3% (Figure S7b).

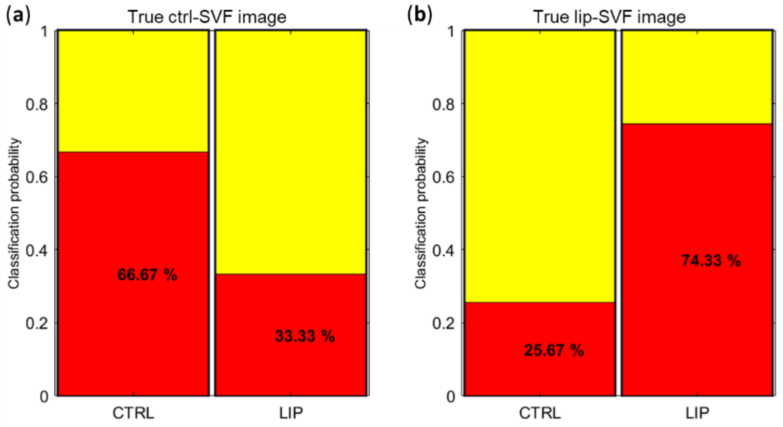


**Figure S7**. Two classification examples of individual SVF-derived EC images. (**a**) Representative result of a ctrl-SVF image. Probability of classification to the ctrl group is 66.67%. (**b**) Representative result of a lip-SVF image. Classification probability to the lip group is 74.33%.

The confusion matrix for this classification is shown in Figure S8. The overall recognition accuracy was 89.7 %. Only six samples (10.3%) were falsely classified, 4 ctrl-SVF images and 2 lip-SVF images.

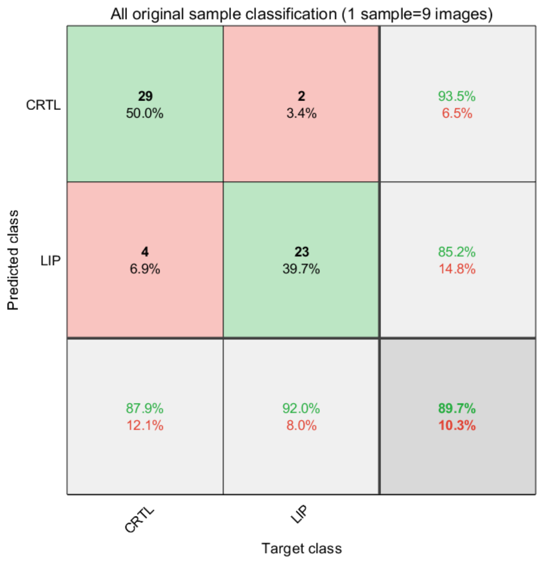


**Figure S8.** Confusion matrix for individual image classification of the SVF-derived EC. 33 samples of ctrl-SVF and 25 samples of lip-SVF were classified by the trained CNN. Recognition accuracy is 89.7%. 6 samples (10.3 %) were falsely classified, 4 ctrl-SVF images and 2 lip-SVF images.

### 3. Experiment 2: CM-treated hEC

To investigate if secreted factors of SVF are able to induce a difference in endothelial junction morphology, hEC were incubated with CM of SVF cells isolated from healthy or lipedema individuals (ctrl-CM and lip-CM, respectively) and immunofluorescence experiments were conducted after 6 days of treatment. 90 images of lip-CM and 120 images of the ctrl-CM were available. From all data, 16 samples from each group were randomly selected as test data. The remaining 74 lip-CM images and 104 ctrl-CM images form the training set. The two groups are not represented equally, therefore the lip-CM group was enlarged with 30 randomly selected duplicates of the original samples, as already described for experiment 1. 9 RGB-images were generated for each sample, which gave a total of 1080 RGB-images per class (with the size of 250 pixel x 250 pixel x 3 channels), 936 in training data set and 144 in data test set. To analyze the reproducibility of the classification, 20 different sets of test images (for 20 independent training and testing sessions) were randomly selected, each consisting of 16 ctrl-CM samples (16 x 9 = 144 images) and 16 lip-CM samples (144 images). The rest of the samples formed the training set. Then, 20 training sessions (separately for each test and training set) of the CNN network with the previously described structure were carried out. The accuracy of the test data classification ranged from 69.8% to 91.01%. The average accuracy of the test data classification for 20 training sessions was 80.2% ± 5.65%. Training data classification approaches ~100 %. The same analysis was performed on a training and test set consisting only of the original images without augmentation (one image per sample i.e. 120 ctrl-CM images and 90 lip-CM images). The average accuracy of the classification for 20 training sessions was 62.7% ± 4.4%. Thus, increasing the number of images by augmenting them significantly increased the accuracy of classification.

An example of training and classification results for one of the test sets is illustrated in Figure S9. Overall classification accuracy of test data was 86.5%. For ctrl-CM images the classification accuracy was 81.9% and for lip-CM images 91.0%. Therefore, images of the ctrl-CM group were more likely to be falsely classified.

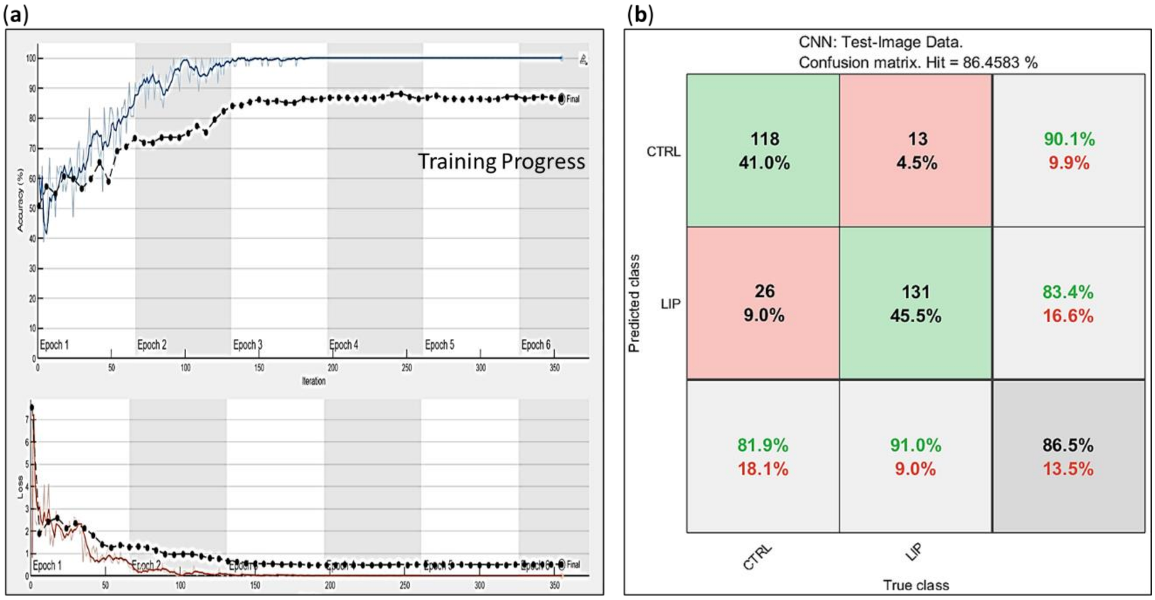


**Figure S9**. Training and classification results for one of the CM-treated hEC test sets (**a**)Training Progress (accuracy in training epochs) and final classification accuracy. (**b**) Confusion matrix for test data set. The columns represent the true classes and the rows the predicted classes. The matrix diagonal gives the absolute and percentage numbers of correctly classified images (green boxes) and the red boxes give the amounts of false classification. The last row of the matrix (grey boxes) gives the cumulative percentage of correct (black letters) and false classifications (red letters) for both classes.

The trained CNN was used as a classifier to predict the belonging of individual samples to a distinct class. Two classification examples are shown in Figure S10. The true ctrl-CM image was classified to the ctrl group with a probability of 88.98 % (Figure S10a), and the true lip-CM image was classified to the lip group with a probability of 83.5% (Figure S10b).

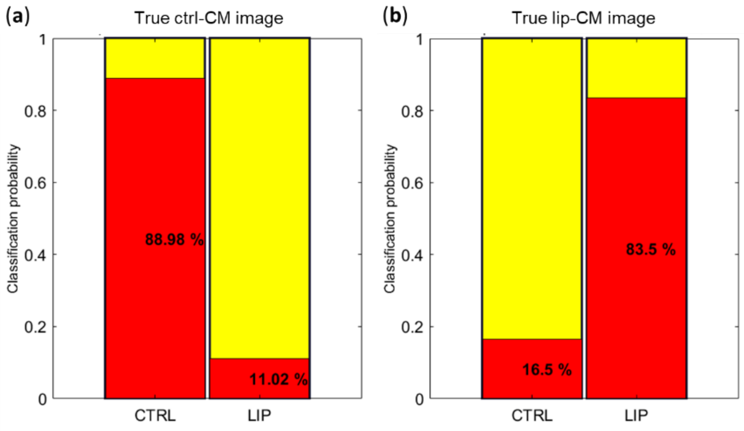


**Figure S10**. Two classification examples of individual images of CM-treated hEC. (**a**) Representative result of a ctrl-CM image. Probability of classification to the ctrl group is 88.98%. (**b**) Representative result of a lip-CM image. Classification probability is 83.5% to the lip group.

Such classification by a trained CNN was performed for total 210 (120 + 90) original samples. The confusion matrix for this classification is shown in Figure S11. The recognition accuracy was ~96%. Only nine images (4.3%) were falsely classified. Training data were classified with an accuracy of ~98%.

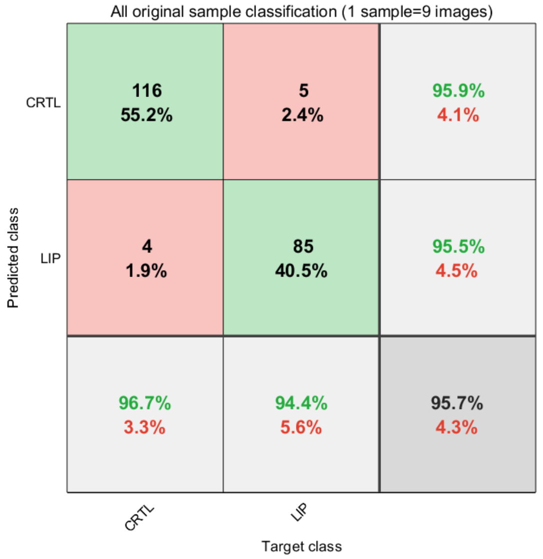


**Figure S11.** Confusion matrix for individual image classification of the CM-treated hEC. 120 samples of ctrl-CM and 90 samples of lip-CM were classified by the trained CNN. Recognition accuracy is 95.7%. 9 samples (4.3%) were falsely classified, 4 ctrl-CM images and 5 lip-CM images.

### 4. Influence of augmented sample-images on classification

Based on the classification of individual samples, we have additionally analyzed which augmented image from a sample (contains 9 images) has the greatest impact on the correct classification. By counting the augmented images for each sample (from the nine-image group), which give the maximum probability of the correct classification, one can build a percentage factor of the image's influence on the classification. The empirical compilation of the impact of individual augmented images is illustrated in Figure S12. Apart from the original, the combined image of the density map, of the segmentation, as well as the skeleton image have the greatest impact on correct classification.

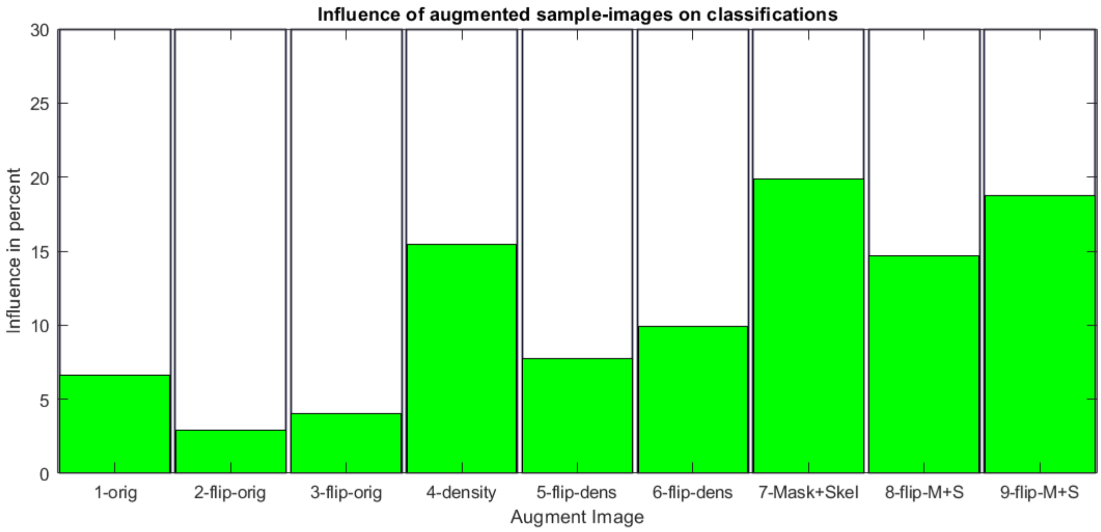


**Figure S12.** Influence of augmented images on correct classification. Abbreviations: 1. Original RGB Image, 2. Flipped up-down original image, 3. Flipped left-right original image, 4. Filtered original image (density map), 5. Horizontal flip filtered image, 6. Vertical flip filtered image, 7. Segmented & skeletonized original image, 8. Horizontal flip segmented & skeletonized image, 9. Vertical flip segmented & skeletonized image.

In summary, the presented CNN neural network classifies all samples at a high level of accuracy. The results confirm that our two-color images as well as their extensions can be used to distinguish the morphology of endothelial junctions of ctrl- from lip-SVF-derived EC, and of hEC treated with lip-CM from ctrl-CM.
